# A liver function test score identifies high-risk MASLD patients based on the pattern of liver enzymes

**DOI:** 10.1101/2024.10.26.24316188

**Authors:** Emma Hajaj, Ahinoam Glusman Bendersky, Marius Braun, Amir Shlomai

## Abstract

**Background & Aims:** A cholestatic pattern of liver enzymes is associated with progressive liver disease and major adverse liver-related outcomes (MALO) among patients with Metabolic Dysfunction-Associated Steatotic Liver Disease (MASLD). We aimed to authenticate the efficacy of a newly formulated liver function test (LFT) score for distinguishing patients with cholestatic vs. hepatocellular patterns and to evaluate its prognostic utility in MASLD patients.

**Methods:** A retrospective longitudinal study on a dataset of over 250,000 individuals diagnosed with MASLD and/or obesity with cardiovascular risk factors. Patients were categorized into cholestatic (C), mixed (M), or hepatocellular (H) patterns according to the LFT score, or the well-known R score. Long-term MALO, major adverse cardiovascular events (MACE), and all-cause mortality were tracked.

**Results:** The LFT score excelled in differentiating patients into C, M, or H groups accurately. While about two-thirds of our cohort initially showed a low FIB4 (<1.3), patients in the C category experienced a higher incidence of MALO and MACE compared to those in the H category (0.5% vs. 0.2% and 7.1% vs. 3.6%, respectively) over the span of 10 years post-diagnosis. Additionally, the 15-year overall survival rate was notably lower for C patients compared to their H counterparts (63% vs. 77%, p<0.0001). The LFT score was more effective than the R score in distinguishing between H and C patients for prognostic purposes, and a baseline cholestatic pattern indicates poorer outcomes regardless of subsequent LFT changes.

**Conclusions:** The LFT score accurately categorizes cholestatic MASLD patients and may serve as a useful prognostic tool.

## Introduction

Metabolic dysfunction-associated steatotic liver disease (MASLD), previously known as non-alcoholic fatty liver disease (NAFLD) (1-3), has a prevalence of ∼30% globally (4, 5). Metabolic dysfunction-associated steatohepatitis (MASH) represents the most severe form of MASLD, posing a significant risk of progressing to advanced liver disease and cirrhosis (4). Patients with MASLD may present with normal liver function tests (LFTs) even in the presence of liver fibrosis (6). However, many MASLD patients present with elevated LFTs that may also have clinical significance; indeed, serum ALT is often used as an indicator for the presence of hepatic steatosis and its progression, and elevated serum ALKP has been linked to advanced MASH (7-9).

Recent studies have indicated that patients with MASLD displaying a cholestatic (C) pattern of liver enzymes tend to have a more severe and progressive form of MASLD. These patients could have worse liver-related long-term outcomes when contrasted with those showing a hepatocellular (H) pattern, which is characterized by raised alanine aminotransferase (ALT) and aspartate aminotransferase (AST), or individuals with a mixed (M) pattern (10-12).

Patients have typically been divided into C or H categories based on the R score, a system originally developed to assess risk in cases of drug-induced liver injury (DILI) (13, 14), which depends only on alkaline phosphatase (ALKP) and ALT levels. In the current study, we introduced an improved LFT score that utilizes all four principal liver enzymes, including gamma-glutamyl transferase (GGT) and AST, which enables finer categorization of MASLD patients according to their profile of elevated LFTs. When we applied this scoring framework to a cohort of over 250,000 patients diagnosed with MASLD, or obesity with cardiovascular risk factors, we observed that individuals categorized with a C pattern by the LFT score (C- LFT) had a significantly increased risk of experiencing major adverse cardiovascular events (MACE), major adverse liver-related outcomes (MALO), and higher overall mortality rates compared to those with an H pattern (H-LFT).

## Materials and methods

### Liver Function Test (LFT) Score

To differentiate between patients with cholestatic (C) or hepatocellular (H) patterns of liver injury, we developed a scoring system using four routinely measured liver enzymes: AST, ALT, ALKP, and GGT. The rationale behind this score was to classify patients based on their liver injury patterns, assigning equal weight to each enzyme. Notably, we included GGT and AST, which are not part of the R score, because ALKP alone is not specific to the liver and cannot serve as the sole indicator of cholestatic liver injury.

To simplify the use of this score and to ensure positive values for hepatocellular injury and negative values for cholestatic injury, the ratio result was transformed using a logarithm. To ensure the values inside the logarithm remained positive and to avoid the issue of taking the logarithm of 0, constants of 2 were added to both the numerator and the denominator. A score exceeding 0.5 indicated a hepatocellular (H) pattern of liver function test (LFT) elevation, while a score below -0.5 indicated a cholestatic (C) pattern. Scores between -0.5 and 0.5 were classified as a mixed (M) pattern of LFT elevation. Patients whose enzyme levels were all within normal ranges at the time of biopsy were excluded from the analysis.

i.e.

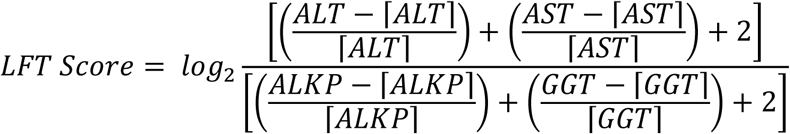

where the names of enzymes denote their serum levels and the enzymes in parentheses (e.g., [ALT]) denote the upper limit of the normal range for each enzyme. The normal ranges used for each enzyme were as follows: AST [0-31 U/L]; ALT [0-34 U/L]; GGT [0-38 U/L]; ALKP [30-120 U/L].

### R score

The R-score was calculated as described in the literature (13) using ALT and ALKP values:

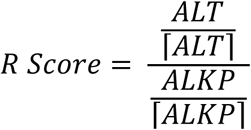

where the names of enzymes denote their serum levels and the enzymes in parentheses (e.g., [ALT]) denote the upper limit of the normal range for each enzyme. The normal ranges for each enzyme were: ALT [0-34 U/L] and ALKP [30-120 U/L].

R-ratios of R>5 define a hepatocellular pattern, R<2 defines a cholestatic pattern, and R between 2 and 5 defines a mixed pattern of enzymes.

### Data from Clalit Healthcare

The analysis of the data from the Clalit Healthcare environment was under IRB approval no. #RMC-23-0322.

Cohort definitions for MASLD – patients with a diagnosis of NAFLD (non-alcoholic fatty liver disease) between the years 2002-2023 were included. Patients with other liver diseases such as HBV, HCV, alcoholic liver disease, autoimmune hepatitis and liver malignancy, as well as liver transplanted patients were excluded.

Cohort definition for obesity and cardiovascular risk – patients with BMI>30 and at least one major cardiovascular risk factor (diabetes mellitus, hypertension, dyslipidemia) between the years 2002-2023 were included. Patients with other liver diseases such as HBV, HCV, alcoholic liver disease, autoimmune hepatitis and liver malignancy, as well as liver transplanted patients were excluded.

For each patient, the LFT and R score were calculated, based on available serum liver enzymes tests at the closest date to the diagnosis of MASLD, as follows: An LFT score exceeding 0.5 indicated a hepatocellular pattern, while a score below -0.5 indicated a cholestatic pattern. As for the R score, R ratio of >5 define a hepatocellular pattern, whereas R<2 defines a cholestatic pattern.

A cohort of 69,513 patients with MASLD, and 193,978 patients with obesity and cardiovascular risk were separated into two groups (hepatocellular vs. cholestatic) according to either the LFT score, or according to the R score.

### Data availability

Due to privacy regulations and organizational policy, all data analysis from Clalit Healthcare Service (CHS) database was conducted on a secured de-identified dedicated server within the Clalit Healthcare environment. Requests for access to all or parts of the Clalit datasets will be considered by the Clalit Research authority, CHS, via a direct request to the corresponding author at. Requests will be considered by CHS research authority upon approval from the institutional review board of CHS and institutional guidelines, and in accordance with the current data sharing guidelines of CHS and Israeli law.

### Statistical analysis

Quantitative variables as lab tests values are expressed as mean ±standard deviation. Categorical variables are expressed as frequencies. Univariate analysis was used to compare baseline characteristics, the mean values of quantitate variables were compared using the independent 2-sided t-test and categorical variables were compared using the chi-square test or Fisher exact test. Statistical significance was set at p value <0.05. Statistical analyses were performed using the GraphPad Prism 10 software.

## Results

### The LFT score classification is more precise than the R score

The LFT score was calculated to equally represent all four liver enzymes, as detailed in the Methods section. Simulated scenarios of various liver enzyme patterns demonstrated that the LFT score more accurately classified patients into the correct group based on their pattern of liver enzyme elevation (Fig 1A). This improved classification is largely due to the low sensitivity of the R score to correctly classify patients with mildly elevated liver enzymes or isolated elevations in GGT or AST, which the R score does not account for.

**Figure 1.**
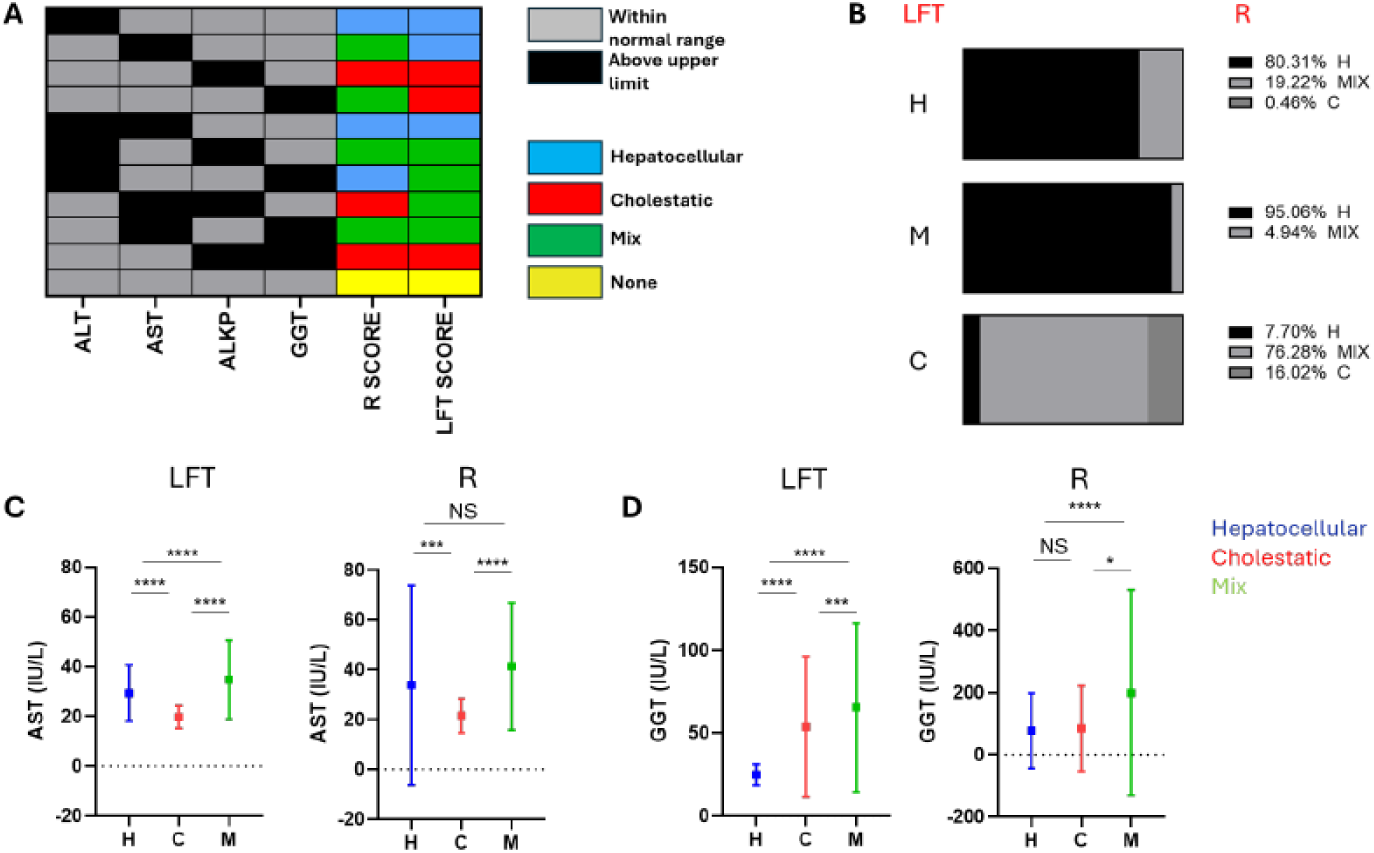
Classification of MASLD patients based on the pattern of their liver enzyme elevations. (A) Graphical presentation of the classification of simulated scenarios of various liver enzyme patterns into hepatocellular (H), cholestatic (C), mixed (M) or no elevation of liver enzymes according to either the R score or the newly computed LFT score. For each case (row) the pattern of liver enzymes (elevated or within normal range) is described according to the color code, and the classification according to the score is depicted. (B) Graphical presentation of the comparison of classification between the two methods (Left – LFT score, right – R score). (C) Mean and standard deviation (STD) of AST levels for MASLD patients classified as cholestatic, hepatocellular or mix according to the LFT score (left) and the R score (right). (D) Mean and STD of GGT levels for MASLD patients classified as cholestatic, hepatocellular or mix according to the LFT score (left) and the R score (right). NS – non significant, * p value<0.05, *** p value<0.001, **** p value <0.0001. two-sided t test.

We next classified 69,513 patients from the Clalit HealthCare system with a documented diagnosis compatible with MASLD, as cholestatic (C), hepatocellular (H), or mixed (M) using both the LFT and the R scores. Comparing the R score and LFT score-based classifications (Fig 1B) revealed that 80.3% of patients classified as H by the LFT score were also predominantly classified as H by the R score, with the remainder mostly classified as M. 95% of patients classified as M by the LFT score were classified as H by the R score, likely due to elevated GGT or mild elevations in ALKP, which the R score does not adequately represent. Notably, only 16% of patients classified as C by the LFT score were similarly classified by the R score, with most being categorized as M. A direct comparison of AST levels across subgroups classified by the LFT and R scores showed the expected low AST levels in the C group for both classifications and higher levels in the H and M groups, with a much wider standard deviation among the H group classified according to the R score (Fig 1C). Notably, GGT levels were significantly higher in the C-LFT and M-LFT groups compared to the H-LFT group (Fig 1D, left panel), whereas in the R classification, GGT levels were similar between the H-R and C-R subgroups and elevated in the M-R subgroup (Fig 1D, right panel).

These findings imply that the LFT score has improved classification accuracy.

### Characteristics of patients as grouped by LFT score and R score classifications

Patients categorized as C by both the LFT and R scores, were predominantly women (70% and 68%, respectively). In contrast, the gender distribution in the H and M groups was more balanced (Table 1). Although most patients across all groups were under 65 years old, those in the C groups were older, with 35% of C-LFT and 28% of C-R patients being over 65, as compared to only ∼18% of patients in the M and H groups.

**Table 1.** Basic demographic parameters of MASLD population according to their classification using the LFT score compared to R score.

|  | LFT |  |  |  |  |  | R |  |  |  |  |  |
| --- | --- | --- | --- | --- | --- | --- | --- | --- | --- | --- | --- | --- |
|  | C | M | H | P value |  |  | C | M | H | P value |  |  |
|  |  |  |  | C vs M | C vs H | M vs H |  |  |  | C vs M | C vs H | M vs H |
| N | 7037 | 4475 | 16765 |  |  |  | 3700 | 6041 | 37986 |  |  |  |
| % | 24.9 | 15.8 | 59.3 |  |  |  | 7.8 | 12.6 | 79.6 |  |  |  |
| Gender (%) |  |  |  |  |  |  |  |  |  |  |  |  |
| Male | 29.9 | 51.6 | 47.71 | **** | **** | **** | 31.46 | 40.13 | 48.17 | **** | **** | **** |
| Female | 70.1 | 48.4 | 52.29 |  |  |  | 68.54 | 59.87 | 51.83 |  |  |  |
| Age (%) |  |  |  |  |  |  |  |  |  |  |  |  |
| Age<65 | 62.33 | 83.22 | 80.42 | **** | **** | 0.04 | 69.35 | 82.85 | 80.08 | **** | **** | 0.016 |
| Age>65 | 34.93 | 16.78 | 17.78 |  |  |  | 28.08 | 17.15 | 18.1 |  |  |  |
| Risk factors 5y before diagnosis (%) |  |  |  |  |  |  |  |  |  |  |  |  |
| DM2 | 39.56 | 35.87 | 26.41 | **** | **** | **** | 29.68 | 24.35 | 30.52 | **** | 0.28 | **** |
| Hypertension | 51.41 | 42.61 | 36.94 | **** | **** | **** | 41.84 | 33.17 | 39.33 | **** | 0.003 | **** |
| Hyperlipidemia | 50.06 | 51.37 | 41.03 | 0.17 | **** | **** | 41.51 | 41.91 | 44.11 | 0.7 | 0.002 | 0.001 |
| Peripheral Vascular Disease | 5.03 | 2.7 | 2.7 | **** | **** | 0.9 | 3.22 | 2.22 | 3.21 | 0.002 | 0.9 | **** |
| FIB4 (% out of samples with relevant data) |  |  |  |  |  |  |  |  |  |  |  |  |
| FIB4<1.3 | 64.69 | 68.6 | 63.61 | **** | **** | **** | 63.01 | 61.73 | 61.43 | **** | **** | 0.001 |
| 1.3<FIB4<2.67 | 31.59 | 20.67 | 30.97 |  |  |  | 31.95 | 27.74 | 30.5 |  |  |  |
| FIB4> 2.67 | 3.72 | 10.73 | 5.42 |  |  |  | 5.04 | 10.53 | 8.07 |  |  |  |

\*\*\*\* - p<0.0001

According to the LFT classification, C-LFT patients exhibited more cardiovascular risk factors and major components of the metabolic syndrome 5 years prior to the diagnosis of MASLD, compared to M-LFT and H-LFT patients, with a higher proportion of patients with type 2 diabetes mellitus (DM2) (39.6% vs. 35.9% and 26.4%, p<0.0001), hypertension (HTN) (51.4% vs. 42.6% and 36.9%, p<0.0001), and peripheral vascular disease (PVD) (5% vs. 2.7% and 2.7%, p<0.0001). Dyslipidemia was equally prevalent in the C-LFT and M-LFT groups, but less common in the H-LFT group (50% and 51%, respectively, vs. 41%, p<0.0001). However, according to the R classification, the difference between C-R and H-R groups was not as significant in the incidence of DM2 (29.7% vs. 30.5%, respectively, p=0.28), HTN (41.8% vs. 39.3%, respectively, p=0.003), and PVD (3.2% in both groups). Interestingly, a higher proportion of both, the C-R and H-R patients, had these conditions compared to M-R patients.

### Patients classified as cholestatic have worse outcomes

We next focused on comparing the main groups: cholestatic (C) vs. hepatocellular (H). In MASLD patients, those classified as C-LFT had lower albumin levels (mean of 4.1g/dL vs. 4.3g/dL respectively, p<0.0001) and prolonged PT-INR (mean of 1.22 vs. 1.1 respectively, p=0.02) compared to H-LFT, with similar trends observed when using the R classification, albeit with a weaker significance (Fig 2A, B). Notably, the predicted level of liver fibrosis, as measured by FIB4, was low (FIB4<1.3) for the majority of patients and did not significantly differ between the groups, regardless of the method of classification (LFT or R) (64.7% vs. 63.6% for C-LFT and H-LFT, respectively, p=0.13 and 63% vs. 61.4% for C-R and H-R, respectively p=0.15) (Table 1).

**Figure 2.**
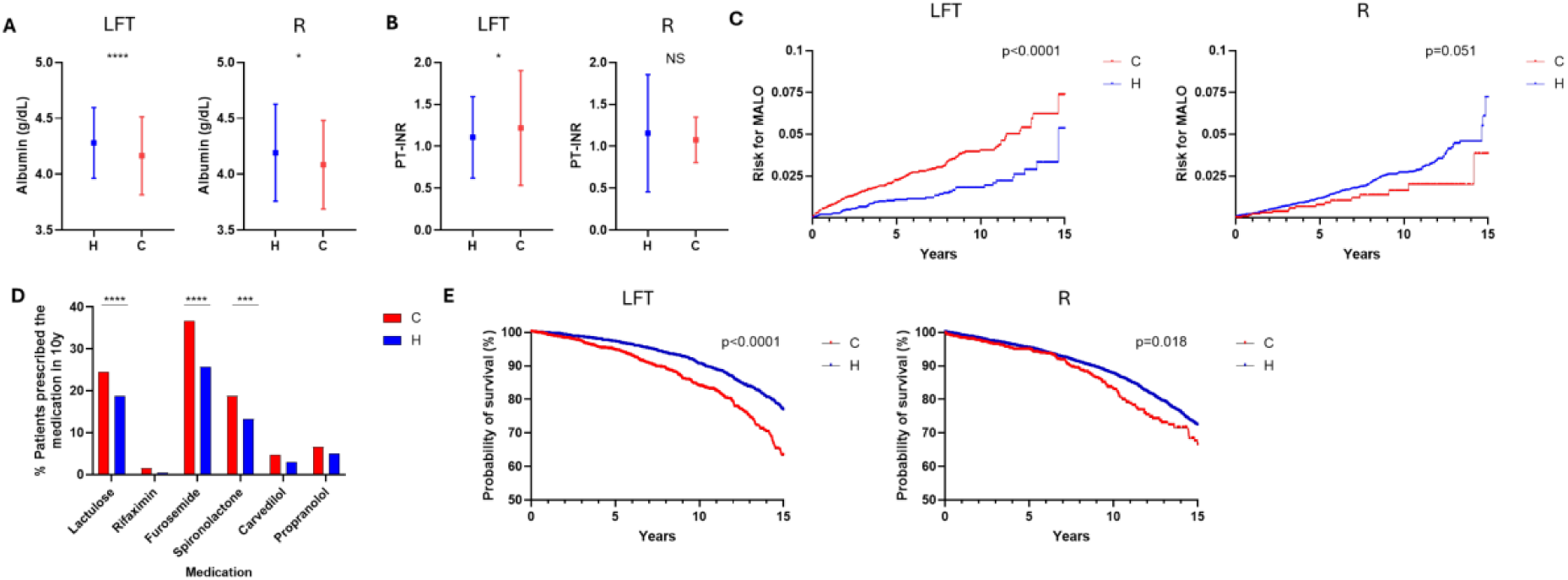
MASLD patients classified as C-LFT have worse prognosis. (A) Mean and STD of albumin levels for MASLD patients classified as cholestatic or hepatocellular according to the LFT score (left) and the R score (right). (B) Mean and STD of PT-INR levels for MASLD patients classified as cholestatic or hepatocellular according to the LFT score (left) and the R score (right). (C) Cumulative risk for major liver related outcomes (MALO) 15 years after diagnosis for MASLD in patients classified as cholestatic or hepatocellular according to the LFT score (left) and the R score (right). (D) Percentage of MASLD patients prescribed medications implicated in the treatment of advanced liver disease during the 10 years post diagnosis, compared between cholestatic and hepatocellular according to LFT score. (E) Kaplan-Meiers survival curves of MASLD patients classified as cholestatic or hepatocellular according to the LFT score (left) and the R score (right). NS – non significant, * p value<0.05, *** p value<0.001, **** p value <0.0001

We further assessed the predictive value of the LFT score for major adverse cardiovascular events (MACE) and major liver-related outcomes (MALO) in a 10-year follow-up. As shown in Table 2, the C-LFT group had a significantly higher incidence of each component of MACE (MI, stroke, heart failure) compared to the H-LFT group, with a higher incidence of each MACE component in the C-LFT group compared to C-R group (6.2% vs. 5.5% for MI, 0.55% vs. 0.38% for stroke, 14.6% vs. 10.6% for heart failure). Overall, as a composite, 7.1% of C- LFT patients had MACE events in 10 years post diagnosis, compared to 3.6% of H-LFT patients.

**Table 2.** Liver related outcomes and MACE in MASLD population according to their classification using the LFT score compared to R score 10 years post diagnosis.

|  | **LFT** |  |  | **R** |  |  |
| --- | --- | --- | --- | --- | --- | --- |
|  | **C** | **H** | **P value** | **C** | **H** | **P value** |
| **N** | 7037 | 16765 |  | 3700 | 37986 |  |
| **MACE (%)** |  |  |  |  |  |  |
| MI | 6.22 | 4.42 | <0.0001 | 5.51 | 4.99 | 0.16 |
| Stroke | 0.55 | 0.35 | 0.034 | 0.38 | 0.47 | 0.52 |
| Heart failure | 14.62 | 6.1 | <0.0001 | 10.57 | 7.91 | <0.0001 |
| **Liver related outcomes (%)** |  |  |  |  |  |  |
| Ascites | 1.63 | 0.55 | <0.0001 | 1.49 | 1.48 | 0.99 |
| SBP | 0.1 | 0.02 | 0.009 | 0.14 | 0.18 | 0.68 |
| Variceal Bleeding | 0.16 | 0.1 | 0.29 | 0.08 | 0.25 | 0.046 |
| Hepatic Encephalopathy | 0.98 | 0.45 | <0.0001 | 0.73 | 0.64 | 0.52 |
| HCC | 0.11 | 0.11 | 0.83 | 0.11 | 0.35 | 0.009 |

The incidence of MALO in 10 years in our cohort was significantly higher in the C-LFT group compared to the H-LFT group (except for variceal bleeding and HCC that did not reach a statistical significance). As a composite, 0.5% of C-LFT patients had MALO events in 10 years, compared to only 0.2% of H-LFT patients. However, these clear differences between the C and H groups regarding MALO were not apparent when using the R classification (Table 2).

The higher cumulative risk for MALO in C-LFT compared to the H-LFT patients (0.075 vs. 0.056 respectively, p<0.0001) was also validated in a 15 year of follow up (Fig 2C, left panel). Conversely, the comparison between the C-R and H-R patients showed only a borderline statistical significance (Fig 2C, right panel).

These differences in MALO incidence were also reflected by the medications prescribed to C- LFT compared to H-LFT patients 10 years after documented diagnosis of MASLD (Fig 2D); C-LFT patients were more frequently prescribed lactulose (24.5% vs. 18.7%) and rifaximin (1.5% vs. 0.5%), diuretics (27.65% vs. 19.5%), and propranolol (6.6% vs. 5%) and carvedilol (4.7% vs. 2.93%), common prescribed medications for hepatic encephalopathy, ascites and varices, respectively.

Given that patients with MASLD and a C-LFT pattern had a greater occurrence of MACE and MALO than those with H-LFT, we next examined whether this disparity affected overall survival rates. Indeed, MASLD patients with a C-LFT pattern have a worse overall survival in 15 years compared to patients with H-LFT pattern (63% vs. 77%, p<0.0001). The variation was likewise noted when contrasting C-R patients with H-R ones, yet the distinction between the cohorts was more marked, and the divergence in their curves occurred sooner when applying the LFT classification than with the R classification (Fig 2E).

### A cholestatic pattern at baseline predicts worse outcomes regardless of future LFT dynamics

Since cirrhotic patients often normalize their LFTs, we next focused on the dynamics of the LFT score over time. As shown in Fig 3A, 65% of MASLD patients classified as C-LFT at diagnosis are still classified the same after 5 years, whereas 30% are classified as M-LFT, and only 4% are classified as H-LFT. In contrast, for patients classified originally as H-LFT, 59% are classified as M-LFT and only 29% are still classified as H-LFT in 5 years.

**Figure 3.**
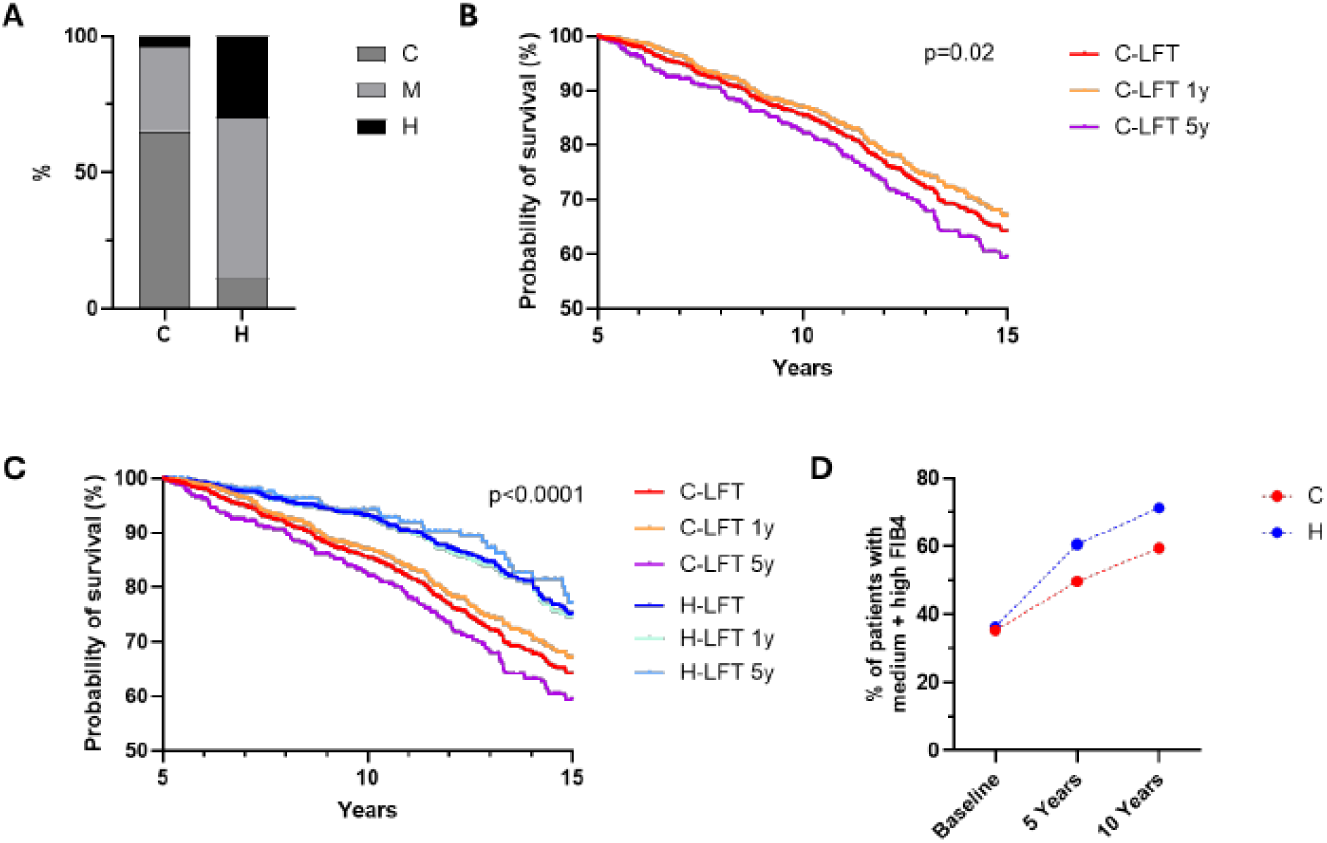
Longitudinal analysis - patients classified as LFT-C at baseline have poor prognosis. (A) Grouping of MASLD patients by LFT score 5 years after diagnosis (shown as fractions in the columns), relative to their initial classification at the time of diagnosis (indicated beside the X axis). (B) A Kaplan-Meier survival curve of all MASLD patients classified as cholestatic at diagnosis (C-LFT), versus patients classified as cholestatic at diagnosis or during the 1^st^ year, and later classified otherwise (C- LFT 1y), and patients that keep the cholestatic classification for 5 years (C- LFT 5y). The p value refers to comparison between the three curves. (C) A Kaplan-Meier survival curve of MASLD patients classified as cholestatic or hepatocellular at diagnosis, patients that kept the classification for 5 years and patients who change their liver enzymes pattern after one year. (D) The percentage of patients with medium (>1.3 but <2.67) or high FIB-4 (>2.67) scores at baseline, 5 years and 10 years post diagnosis, according to their baseline C or H pattern.

In a 15 year of follow-up, patients with persistence C-LFT pattern within 5 years from diagnosis have a worse overall survival compared to patients with persistence C-LFT during the first year after diagnosis who switched later on to another category (59.4% vs. 67%, p=0.02) (Fig 3B). Most important, patients with C-LFT pattern at diagnosis who switched their pattern after the first year following diagnosis, still have a much worse long-term prognosis compared to patients with H-LFT pattern at diagnosis, regardless of whether they maintained the H status at 5 years or only during the first year following diagnosis (Fig 3C).

We next aimed to evaluate changes in the FIB4 score, a non-invasive liver fibrosis marker, over time in our cohort of MASLD patients. Surprisingly, those with H-LFT at baseline had a higher percentage of patients with FIB4>1.3 at 5- and 10-years post-diagnosis compared to those with C-LFT at baseline (Fig. 3D), although most of the H-LFT patients with high FIB4 at 10 years were still at the H-LFT category, and only 12.6% were at the C-LFT category according to their lab tests at 10 years.

These results suggest that the pattern of liver enzymes at baseline among MASLD patients may have an impact on long-term prognosis, regardless of its future dynamics, and that the Fib4 dynamics overtime may underestimate the severity of the disease in MASLD patients with a cholestatic pattern.

### LFT score in obese patients with cardiovascular risk factors

To reduce potential misclassification bias of MASLD in this large dataset, we employed the LFT and R scores on a cohort of 193,978 obese individuals with at least one major cardiovascular risk factor (type 2 diabetes, hypertension, or dyslipidemia) who had no other documented liver diseases. This group of patients is at a very high-risk for MASLD. Indeed, there was a considerable overlap between the groups; 16% of these individuals had a formal diagnosis that aligned with MASLD and were consequently part of the previously mentioned MASLD cohort, and out of the MASLD cohort, 45% were included in the obesity cohort. Table 3 summarizes the characteristics of these patients according to both, LFT- and R-based classifications. The gender and age distributions were consistent with those observed in the MASLD cohort.

**Table 3.** Basic demographic parameters of obese patients with cardiovascular risk factors according to their classification using the LFT score compared to R score.

|  | LFT |  |  |  |  |  | R |  |  |  |  |  |
| --- | --- | --- | --- | --- | --- | --- | --- | --- | --- | --- | --- | --- |
|  | C | M | H | P value |  |  | C | M | H | P value |  |  |
|  |  |  |  | C vs M | C vs H | M vs H |  |  |  | C vs M | C vs H | M vs H |
| N | 25703 | 15109 | 34509 |  |  |  | 18032 | 33247 | 66188 |  |  |  |
| % | 34.12 | 20.05 | 45.8 |  |  |  | 15.35 | 28.3 | 56.3 |  |  |  |
| Gender (%) |  |  |  |  |  |  |  |  |  |  |  |  |
| Male | 38.9 | 56.5 | 55.5 | **** | **** | 0.04 | 38.84 | 47.71 | 55.62 | **** | **** | **** |
| Female | 61.1 | 43.5 | 44.5 |  |  |  | 61.16 | 52.29 | 44.38 |  |  |  |
| Age (%) |  |  |  |  |  |  |  |  |  |  |  |  |
| Age<65 | 47.01 | 57.03 | 62.55 | **** | **** | **** | 60.93 | 68.92 | 65.44 | **** | **** | **** |
| Age>65 | 50.24 | 40.75 | 34.86 |  |  |  | 39.07 | 31.08 | 34.56 |  |  |  |
| FIB4 (% out of samples with relevant data) |  |  |  |  |  |  |  |  |  |  |  |  |
| FIB4<1.3 | 57.9 | 41.63 | 46.66 | **** | **** | **** | 61.15 | 49.05 | 47.57 | **** | **** | **** |
| 1.3<FIB4<2.67 | 35.86 | 37.22 | 37.8 |  |  |  | 31.48 | 30.48 | 36.1 |  |  |  |
| FIB4> 2.67 | 6.24 | 21.15 | 15.53 |  |  |  | 7.37 | 20.48 | 16.33 |  |  |  |

Interestingly, a higher proportion C patients had a low FIB4 (<1.3) compared to H and M patients in this population, regardless of the classification method used.

Despite this, and similar to the documented MASLD population, liver synthetic function tended to be worse in C-LFT compared to H-LFT patients, with prolonged INR and lower albumin values, in the formers (Fig 4A, B).

**Figure 4.**
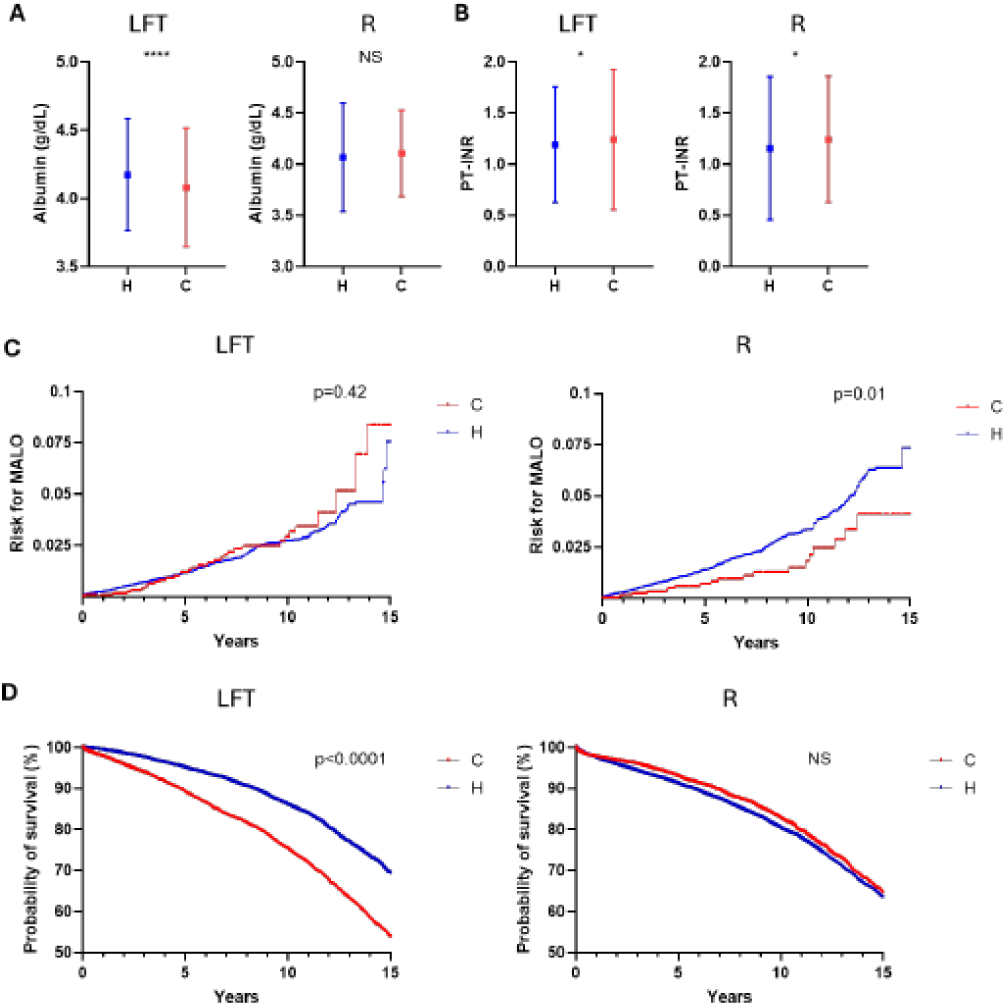
Obese patients with CV risk factors classified as LFT-C have worse prognosis. (A) Mean and STD of Albumin levels for obese patients classified as cholestatic or hepatocellular according to the LFT score (left) and the R score (right). (B) Mean and STD of PT-INR levels for obese patients classified as cholestatic or hepatocellular according to the LFT score (left) and the R score (right). (C) Cumulative risk for liver related outcomes 15 years after diagnosis for obese patients classified as cholestatic or hepatocellular according to the LFT score (left) and the R score (right). (D) Kaplan-Meiers survival curves of obese patients classified as cholestatic or hepatocellular according to the LFT score (left) and the R score (right). NS – non significant, * p value<0.05, **** p value <0.0001

Among this cohort, the H-LFT group had a higher incidence of myocardial infarction (MI) compared to the C-LFT group (13.8% vs. 12.2%, p<0.0001), while other components of MACE were significantly more frequent in the C-LFT group. However, according to the R classification, the C-R group did not have a higher frequency of any component of MACE compared to the H-R group (Table 4).

**Table 4.** Liver related outcomes and MACE in obese patients with cardiovascular risk factors according to their classification using the LFT score compared to R score 10 years post diagnosis.

|  | **LFT** |  |  | **R** |  |  |
| --- | --- | --- | --- | --- | --- | --- |
|  | **C** | **H** | **P value** | **C** | **H** | **P value** |
| **N** | 25703 | 34509 |  | 18032 | 66188 |  |
| **MACE (%)** |  |  |  |  |  |  |
| MI | 12.29 | 13.84 | <0.0001 | 9.59 | 13.73 | <0.0001 |
| Stroke | 1.07 | 0.82 | 0.0012 | 0.87 | 0.85 | 0.89 |
| Heart failure | 27.48 | 17.08 | <0.0001 | 17.32 | 19.49 | <0.0001 |
| **Liver related outcomes (%)** |  |  |  |  |  |  |
| Ascites | 1.86 | 1.08 | <0.0001 | 1.22 | 1.82 | <0.0001 |
| SBP | 0.08 | 0.03 | 0.03 | 0.06 | 0.13 | 0.006 |
| Variceal Bleeding | 0.04 | 0.08 | 0.048 | 0.05 | 0.17 | <0.0001 |
| Hepatic Encephalopathy | 0.93 | 0.59 | <0.0001 | 0.65 | 0.87 | 0.0035 |
| HCC | 0.19 | 0.17 | 0.55 | 0.14 | 0.38 | <0.0001 |

In addition, C-LFT patients had a higher incidence of ascites, hepatic encephalopathy, SBP and variceal bleeding compared to H-LFT in a 10 year follow-up (Table 4). However, using the R classification, all liver-related outcomes were significantly more common in the H-R group compared to the C-R group.

Analysis of the cumulative risk for MALO in a 15 year follow-up showed a trend towards a higher incidence in C-LFT compared to H-LFT patients, that did not reach a statistical significance. However, the opposite was observed in C-R compared to H-R patients (Figure 4C).

Most important, C-LFT patients had a worse overall survival in 15 years compared to H-LFT patients (54% vs. 69.3%, p<0.0001), whereas this difference was not observed between C-R compared to H-R patients (Figure 4D).

## Discussion

MASLD is emerging as the fastest growing and the leading etiology for liver disease in many countries around the world (15). While most individuals diagnosed with MASLD do not develop severe liver disease, they are at an increased risk for conditions associated with the metabolic syndrome and cardiovascular diseases (16). Still, liver fibrosis is associated with increased risk of mortality and liver-related outcomes among patients with MASLD (17), and therefore there is much interest in non-invasive tests (NITs) for early detection of significant liver fibrosis and allocation of high-risk MASLD patients(18, 19).

Recent findings indicate that patients with MASLD with a cholestatic pattern of liver enzymes tend to experience more aggressive disease progression that might also correlate with a distinct pattern of liver histology(20, 21). However, these conclusions are mainly based on the R score that incorporates only ALT and ALKP levels, as the basis for categorization of patients’ liver enzymes as cholestatic or hepatocellular. This might lead to inaccurate classification of patients who have mainly GGT increases, as sometimes seen in the metabolic syndrome(22, 23), or those with standalone ALKP increases that may not stem from hepatic causes.

In this study, we utilized a newly developed LFT score that incorporates all four major liver enzymes (ALT, AST, ALKP, GGT) routinely measured in clinical practice, to define MASLD patients as having either a cholestatic (C) or hepatocellular (H) pattern of liver enzymes abnormalities. We show that in comparison to the R score, the LFT score more accurately categorizes patients as having H, M or C patterns of liver enzymes. This is also evident in the study of Pennisi et al. (11) involving a large cohort of MASLD patients categorized on the basis of the R score, where GGT level did not significantly differ between the C, M or H groups.

In agreement with our previous report(10), the current study shows that MASLD patients (or patients with obesity with at least one additional cardiovascular risk factor) with a cholestatic pattern, are older with a tendency for a female predominance. More importantly, and consistent with previous reports(10, 11), our findings indicate that C patients exhibit a higher prevalence of baseline CV risk factors than those with H pattern, with a more distinct difference observed using the LFT score rather than the R score. Correspondingly, the incidence of MACE over a decade was notably higher in the C group compared to the M and H groups, with the LFT score providing a clearer distinction between the high-risk C group and the other H and M groups.

Consistent with the study of Pennisi et al.(11), our results strongly suggest that a cholestatic pattern in MASLD, as well as in high-risk patients having obesity and at least one major CV risk factor, is associated with an increased risk for MALO in a follow-up of up to 15 years. In our study, this distinction is clear upon categorization of MASLD patients according to the LFT score and less so with the R score. Furthermore, among patients with obesity and at least one CV risk factor, this difference is seen only when using the LFT score to categorize these high-risk patients into C or H groups.

One might hypothesize that patients with MASLD exhibiting a C pattern could be more prone to developing advanced liver fibrosis or cirrhosis, as suggested by previous reports (10, 11), highlighting a connection between the C pattern and an increased risk of later developing MALO. Regrettably, our comprehensive study of MASLD and metabolically high-risk patients does not include data on liver histology. Nevertheless, in our MASLD patient group, the distribution of FIB4 scores was largely consistent across H, M, and C categories, with roughly two-thirds having a low FIB4 score (<1.3) in each category at baseline. Interestingly, a significantly higher percentage of H-LFT patients were in the Fib4>1.3 category than C-LFT patients at both 5 and 10-year follow-ups. Furthermore, in obese patients with cardiovascular risk factors, fewer than half of those with M or H patterns had low FIB4 scores, while about two-thirds of those with a C pattern did. These unexpected findings might highlight the diminished effectiveness of the FIB4 score, incorporating hepatocellular rather than cholestatic liver enzymes, in detecting patients with a C pattern who have advanced liver fibrosis. FIB4, now adopted by international guidelines as a simple non-invasive screening test for advanced liver fibrosis in high-risk MASLD patients(24, 25), has been shown to have a prognostic utility for liver and cardiovascular events, as well as for overall mortality in patients with MASLD(26, 27). Further studies should better define the role of FIB4 as a predictor for advanced liver fibrosis and MALO in this subset of patients who present with a cholestatic pattern of liver enzymes. Indeed, although non-invasive tests such as FIB4 has a considerable accuracy in predicting advanced fibrosis in MASLD patients(28), recent reports point out to a discordance between the FIB4 score and liver fibrosis, as evaluated by transient elastography, in certain populations of MASLD patients who are mainly older with more severe metabolic derangements and higher ALT, AST and ALKP levels(29).

Our study shows that among MASLD and high-risk obese patients, the LFT score outperforms the R score in predicting long-term survival that is worse for C compared to H patients. In addition, our study presents a noteworthy finding regarding the temporal behavior of liver enzyme patterns and their prognostic significance; The vast majority of MASLD patients with C pattern at diagnosis remained at the same category after 5 years, and this group of patients have a worse overall survival compared to patients who remain at this category only for the first year following their diagnosis. In contrast, only about 30% of H-LFT patients at diagnosis remain at their “benign” category after 5 years. However, this group of patients still have a better prognosis compared to patients initially categorized as C, even if switched to another more “benign” category after the first year from diagnosis. While the exact mechanism behind this finding is uncertain, it indicates the potential value of tracking changes in LFT scores, possibly with conjunction of other non-invasive tests, longitudinally.

The strength of our study is its large cohort of patients for whom a vast amount of documented data was available, and its longitudinal design. Its main limitations are its retrospective nature and the lack of data regarding liver fibrosis status documented by either liver biopsy or by other non-invasive methods such as transient elastography. Given our dependence on medical records and to reduce the possibility of incorrectly identifying patients with MASLD, we also conducted a comparable study on a sizeable group of obese individuals who have at least one other major cardiovascular risk factor. This group is likely to suffer from MASLD(30), and indeed partially overlapped with our MASLD cohort, and contemporary screening recommendations for advanced liver fibrosis apply to them.

In summary, our results suggest that LFT score can categorize MASLD patients into C or H patterns and that that C pattern is implicated in a greater risk for MALO, MACE and worse long-term overall survival. The LFT score outperforms the R score in categorization into a distinct pattern of liver enzymes. Future research may investigate the predictive value of the LFT score in conjunction with commonly used non-invasive tests like the FIB4 among different groups of MASLD patients.

## Abbreviations

MALO: major adverse liver-related outcomes
MASLD: Metabolic Dysfunction- Associated Steatotic Liver Disease
LFT: liver function test
MACE: major adverse cardiovascular events
MASH: Metabolic dysfunction-associated steatohepatitis
ALT: alanine aminotransferase
AST: aspartate aminotransferase
ALKP: alkaline phosphatase
GGT: gamma-glutamyl transferase

## Authors’ contributions

Emma Hajaj- Conceived and designed the analysis, collected and analyzed the data, wrote the manuscript draft

Ahinoam Glusman Bendersky- Statistical analysis

Marius Braun- Contributed data and logistic tools

Amir Shlomai- Conceptualization, conceived and designed the analysis, wrote and edited draft and final manuscript, supervised the project

## Ethics and data availability statements

(see Methods section)

